# Sustained Viral Suppression and its association with Psychiatric Morbidity and Sleep Quality among Older Adults with Human Immunodeficiency Virus: Evidence from a Retrospective Cohort in Ibadan, Nigeria

**DOI:** 10.64898/2026.10.07.26364957

**Authors:** Olufisayo Elugbadebo, Faithful Akintade, Bibilola Oladeji, Lauren Brittany Beach, Lisa M. Kuhns, Chiamaka Egbuna, Olutosin Awolude, Olayinka Omigbodun, Philip B. Gorelick

**Author notes:** **Corresponding Author:** Name: Philip B. Gorelick, Adress: Davee Department of Neurology and Havey Institute for Global Health, Northwestern University Feinberg School of Medicine.

## Abstract

**Purpose:** Older adults with HIV are at risk of psychiatric disorders and poor sleep quality, but evidence from sub-Saharan Africa is limited, particularly regarding sustained viral suppression. This study examined the prevalence of psychiatric disorders and poor sleep quality symptoms and their association with sustained viral suppression among older adults with HIV in Nigeria.

**Methods:** A retrospective cohort study was conducted among older adults receiving care at a tertiary HIV clinic in Ibadan, Nigeria. Viral load data from 2019-2023 were abstracted from medical records to determine sustained viral load suppression, while psychiatric morbidity and sleep quality were assessed cross-sectionally in 2023. Psychiatric morbidity, including major depressive disorder, anxiety disorder, bipolar disorder and mania, was assessed using the World Mental Health Composite International Diagnostic Interview (WMH-CIDI), while sleep quality was assessed using the Pittsburgh Sleep Quality Index (PSQI). Mixed-effects logistic regression assessed changes in viral suppression over time and associations with psychiatric morbidity and sleep quality.

**Results:** Among 400 participants, 63.5% were female and mean age was 58.2 years (± 6.5). The 12-month prevalence of major depressive disorder was 25.8% while 17.5% had anxiety disorder, 29.3% had poor sleep quality, 3.3%, bipolar disorder, and 1.0% mania. Viral suppression improved significantly over the retrospective period. Sustained viral suppression was significantly associated with higher odds of good sleep quality (OR = 1.69, 95% CI: 1.07–2.66, p = 0.025), but not psychiatric morbidity.

**Conclusion:** Psychiatric disorders and poor sleep quality are common among older adults with HIV in Nigeria. Sustained viral suppression was associated with better sleep quality but not psychiatric morbidity highlighting the importance of integrating mental health and sleep assessment into comprehensive HIV care for older adults.

## Introduction

Advances in antiretroviral therapy (ART) have transformed HIV infection into a chronic condition, leading to marked improvements in survival globally[1–3], Consequently, a growing population of people with HIV (PLWH) are now aged 50 years and above, representing a major public health success of ART[1, 4–6]. Although the overall number of People Living with HIV (PLHIV) in Nigeria has remained relatively stable in recent years, the population of older PLHIV continues to grow. As of 2024, an estimated 480,000 (350,000–710,000) people aged 50 years and older were living with HIV in Nigeria, representing approximately 24% of the country’s estimated 2.0 million people with HIV[7]. This reflects the increasing population-level aging of PLHIV, following expanded access to effective treatment.[8].

HIV treatment success has traditionally been assessed using biomedical indicators, particularly viral suppression, as reflected in the UNAIDS 95–95–95 targets[9]. However, these measures provide limited information about quality of life and mental well-being, especially among older adults. This has informed increasing emphasis on the “fourth 95,” focused on mental health and functional outcomes[10].

Mental health remains weakly integrated into HIV services for older adults in many low- and middle-income settings[11]. Older Persons Living with HIV (OPLHIV) experience higher rates of mental disorders and cognitive impairment[12, 13], reflecting the cumulative effects of aging, chronic HIV infection, long-term ART exposure, and sustained psychosocial stressors, including stigma, social isolation, multimorbidity, and limited access to mental health services[14–17]. In many Nigerian HIV clinics, mental health assessment is largely symptom-driven, potentially contributing to under-recognition of depression and poor sleep quality, thereby limiting timely diagnosis and appropriate management of these comorbidities. Psychiatric morbidity among PLWH has also been associated with poorer ART adherence and reduced engagement in care[18, 19]. Despite this, HIV outcomes continue to be assessed largely through viral suppression and preserved immune function [20]. Early immune damage, reflected by baseline CD4 count, and viral suppression over time have been hypothesized to be associated to relate to psychiatry morbidity including depressive symptoms.[21]. However, this relationship remains insufficiently examined among OPLHIV.

Depression is consistently associated with poorer ART adherence, while poor adherence may also contribute to psychological distress through treatment failure, worsening physical health, concerns about disease progression[22]. Both may also share psychosocial determinants, such as HIV-related stigma, limited social support[23], and poor socioeconomic conditions[24]. These mechanisms suggest a potentially reciprocal relationship.

Understanding how psychiatric morbidity relates to virologic indicators may help clarify whether conventional markers of HIV treatment success (viral load) adequately reflect overall health in later life. Sleep quality is also relevant because good sleep is associated with better physical health, mood, functioning, and quality of life. [25]. In PLHIV, poor sleep quality may interfere with treatment adherence. Furthermore, poor sleep quality may interfere with glymphatic clearance of toxic waste in the brain which occurs largely during sleep.

In Nigeria, HIV research has largely focused on younger populations, emphasizing behavioral outcomes and access to treatment[26]. Evidence on psychiatric morbidity and poor sleep quality among OPLHIV and their relationship with viral suppression overtime, remains limited[27]. In this context, this study aimed to determine the prevalence of major depressive disorder, anxiety disorders and poor sleep quality symptoms among OPLHIV. It also sought to assess the proportion of participants with major depressive disorder or anxiety disorders who had ever received biomedical treatment, and to examine the association of psychiatric morbidity and sleep quality with sustained viral suppression.

## Methodology

### Study design

This study used a retrospective cohort study design to examine longitudinal viral suppression among OPLHIV aged ≥50 years with psychiatric morbidity and sleep quality assessed cross-sectionally in 2023 at a tertiary HIV care clinic in Ibadan, Nigeria. A retrospective cohort design was selected to allow measurement of sustained viral suppression across time prior to the collection of psychiatric morbidity and sleep quality data, which were assessed using validated research instruments administered by trained research staff. The Pittsburgh Sleep Quality Index (PSQI) assessed self-reported sleep quality over the preceding month, whereas the Composite International Diagnostic Interview (CIDI) established 12-month diagnoses of major depressive and anxiety disorders.

### Study setting

The study was conducted at the AIDS Prevention in Nigeria (APIN) clinic, Infectious Disease Institute of the College of Medicine, University of Ibadan (IDI/CoMUI). The clinic is supported by the U.S. President’s Emergency Plan for AIDS Relief (PEPFAR) and provides HIV screening and treatment services to patients referred from all levels of healthcare facilities within the region and its environs. Eligible patients receive free antiretroviral therapy (ART) with scheduled pharmacy visits, and medications refills are routinely documented. The clinic electronic database contains demographic and longitudinal clinical information, including ART and other medications, baseline CD4 cell counts, and annual viral load measurements. As psychiatric morbidity is not routinely assessed in the clinic, mental health and sleep outcomes were assessed separately in 2023 using standardized instruments.

### Study participants

Participants were OPLHIV aged ≥50 years who received care at the clinic between 2019 and 2023 and were recruited consecutively during routine clinic visits and included in the analysis if eligible. Eligibility criteria included: (a) age ≥50 years; (b) receiving care at the IDI; (c) ART use from 2019 to 2023. Participants were excluded if: (a) unable to provide informed consent or assent; (b) had acute medical illness requiring hospitalization; or (c) had severe cognitive impairment (MoCA score <10). All eligible participants provided informed consent and were informed of their right to withdraw without consequence. Data were de-identified using unique study identifiers distinct from clinic records and stored securely with access restricted to authorized study personnel. Participants received reimbursement for transportation costs.

### Variables

#### Data sources and measurement

Sociodemographic, mental health, and sleep quality data were collected from participants at the point of enrollment, while clinical variables were concurrently abstracted from their medical record. Viral load measurements were retrospectively extracted from medical records over the preceding 4-year period to determine sustained viral load suppression.

Primary outcomes, including major depressive disorder, anxiety disorders and sleep quality, were assessed using validated instruments (CIDI and PSQI) administered by trained research assistants. Clinical variables including CD4 cell count and annual HIV viral load were obtained from electronic medical records and laboratory databases. Demographic and clinical factors, including age, sex at birth, education, year of HIV diagnosis, and duration of ART were obtained using interviewer administered questionnaires.

### Outcome measures

#### The World Mental Health Composite International Diagnostic Interview (WHO WMH-CIDI)

The WHO WMH-CIDI was used to assess past 12-month prevalence of major depressive disorder, anxiety disorders, bipolar disorder and mania, as well as previous professional treatment for major depression and anxiety. The CIDI is a comprehensive, fully structured interview for assessing mental disorders according to ICD-10 and DSM-IV criteria[28]. Responses were analyzed using the standard WHO WMH-CIDI diagnostic algorithms to establish 12-month diagnoses of major depressive disorder and anxiety disorder according to the ICD10 criteria (Appendix).

#### Sleep Quality

Sleep quality was evaluated using the PSQI, a validated instrument measuring sleep quality and disturbances over the preceding month. The seven component scores generate a global score ranging from 0 to 21, with higher scores reflecting poorer sleep quality. A global PSQI score >5 was used to define poor sleep quality[29].

#### Bias

Potential bias was minimized using validated CIDI and PSQI and through administration of the study instruments by trained interviewers. Consecutive recruitment of eligible participants was carried out to reduce selection subject selection bias, and consistent assessment methods were applied across all participants.

#### Sample size

The study population comprised OPLHIV (≥50 years) receiving ART at IDI. As of November 2022, a total of 2,851 eligible individuals were receiving care. The sample size was determined a priori using Yamane’s formula [30] for a finite population assuming a 95% confidence level and a 5% margin of error. The minimum required sample size was calculated as 351 participants, allowing 10% for non-response or incomplete data, yielded 386 participants, which was rounded up to a target of 400 participants.

#### Data collection and measurements

Sociodemographic and clinical data included age, sex at birth, CD4 cell count and viral load measurements., Viral load measurements from 2019 to 2023 and baseline CD4 cell count were obtained from clinical records. Psychiatric morbidity, including depression, anxiety, bipolar disorder, and mania, was assessed using the WHO WMH-CIDI while sleep quality was assessed using PSQI. Distributions were assessed for normality using histograms and summary statistics; and log-transformed where appropriate.

##### Variable definition for analysis

Age was summarised both as a continuous variable (mean and standard deviation) and as a categorical variable in the descriptive analyses but retained as a continuous variable in inferential analyses. Baseline CD4 cell count was described according to the CDC’s staging system[31]: stage 1: CD4 count≥500 cells/μL stage 2: CD4 count 200-500 cells/μL stage 3: CD4 count ≤ 200 cells/μL (CD4 count was retained as a continuous variable in the regression analysis

Viral suppression was defined using the WHO threshold of ≤1000 copies/mL = viral suppression and >1000 copies/mL = unsuppressed. Using all available measurements from 2019 - 2023, sustained viral suppression was defined as viral load of ≤1,000 copies/mL at all available time points, whereas unsustained viral suppression was defined as at least one measurement >1,000 copies/mL. Sustained viral suppression was coded as 1 and unsustained viral suppression as 0. Depression, anxiety, bipolar disorder and mania were classified according to CIDI diagnostic algorithms and coded as binary variables (1 = diagnosis present; 0 = diagnosis absent). A PSQI >5 indicated poor sleep quality. Sleep quality was dichotomized as good sleep (coded 1) and poor sleep (coded 0).

#### Statistical Methods

Participant characteristics were summarized using means and standard deviations for continuous variables and frequencies and percentages for categorical variables. Odds ratios and 95% confidence intervals (CI) were used to estimate the strength of associations between the independent variables and categorical outcomes.

#### Analysis of viral suppression over time

Changes in annual viral suppression from 2019-2023 were examined using generalized estimating equations (GEE) model with a binomial distribution and logit link, with participant ID specified as the subject variable to account for repeated observation.[32] An exchangeable working correlation structure was specified, assuming constant correlation between repeated measurements from the same individual across study years. GEE was selected because it estimates population-averaged effects and accommodates unbalanced longitudinal data. Therefore, participants with missing viral load measurements for one or more study years were retained in the analysis and contributed all available observations, rather than being excluded.[33]. The model used 2019 as the reference year, with indicator variables for 2020, 2021, 2022, and 2023 to estimate year-specific differences in odds of viral suppression. Year was subsequently modelled as a continuous variable to evaluate the overall linear trend in viral suppression from 2019-2023.

#### Analysis of sustained viral suppression

Major depressive disorder, anxiety disorders, and sleep quality assessed in 2023 were treated as binary outcomes. Their associations with sustained viral load suppression (2019-2023), baseline CD4 cell count, and sociodemographic factors were assessed using binary logistic regression. Due to the small number of mania (n = 4) and bipolar cases (n = 13), these outcome were excluded from multivariable regression[34]to avoid overfitting and unstable estimates, and were reported descriptively only.

Statistical significance was set at a two-sided p-value <0.05. Analyses were performed using SPSS version 27.

**Fig 1:**
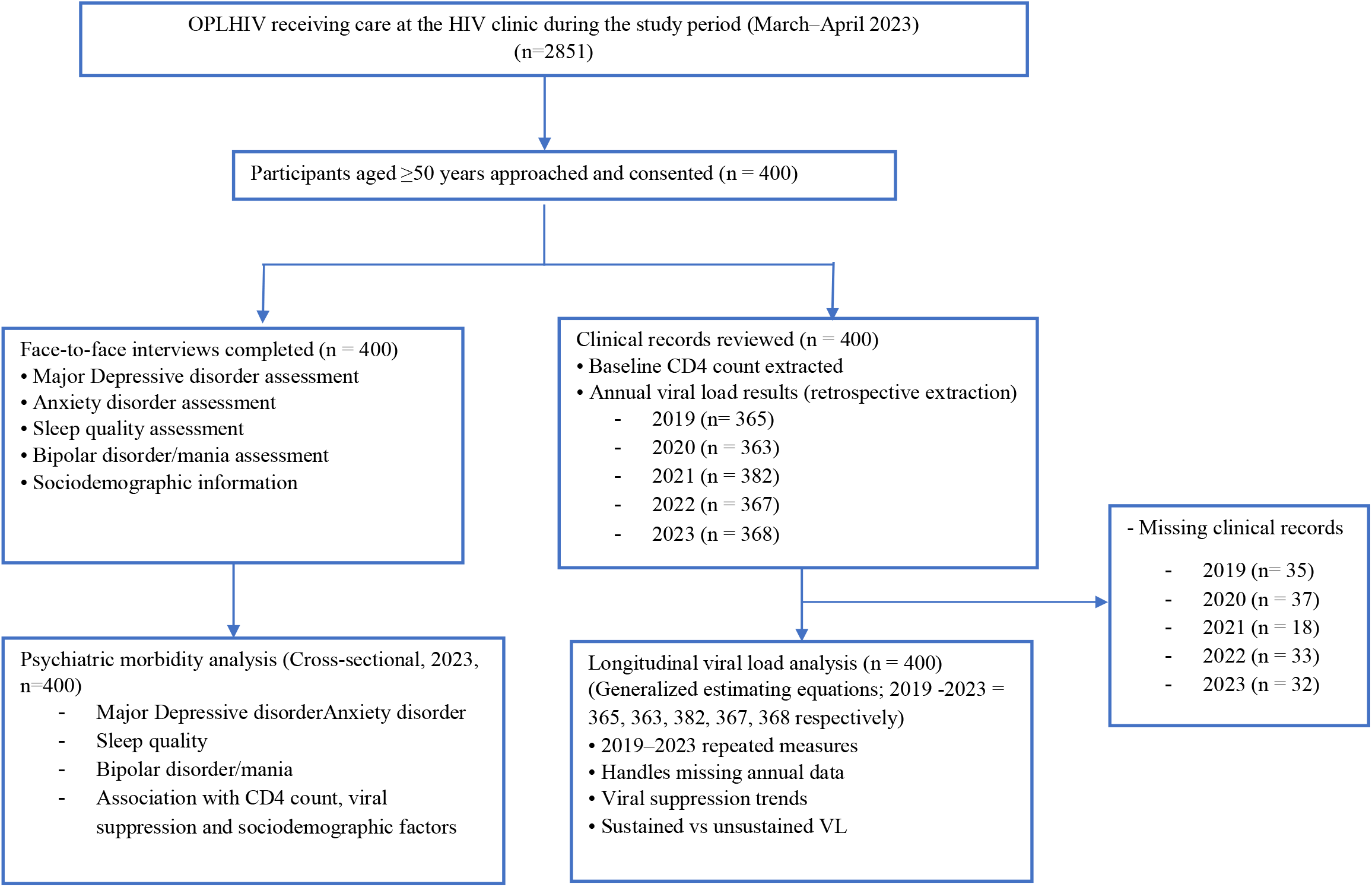
STROBE flow diagram showing participant inclusion and availability of longitudinal clinical and mental health data

## Results

### Characteristics of the Participants

Of the 400 participants included in this study, mean age was 58.2 years (SD = 6.5 years), with the majority (63.5%) being female. In terms of education, the largest percentage, 36.5%, obtained tertiary education, most were currently married (55.0%), were Christian (69.2%), and were living with others (81.5%). In terms of clinical history, most were diagnosed with HIV between 2005 and 2012 (68.7%), and 58.3% were age 38-48 years at the time of diagnosis. (Table 1).

**Table 1:** Socio-demographic Characteristics of the Study participants (n=400)

| ***Variables*** | ***Frequency*** | ***Percentage (%)*** |
|---|---|---|
| ***Current Age*** |  |  |
| *50-59* | 257 | 64.3 |
| *60-69* | 115 | 28.7 |
| *70 and above* | 28 | 7.0 |
| *Mean (S.D)* | 58.2 | 6.5 |
| ***Sex at birth*** |  |  |
| *Male* | 146 | 36.5 |
| *Female* | 254 | 63.5 |
| ***Educational Attainment*** |  |  |
| *No formal Education* | 24 | 6.0 |
| *Primary education* | 95 | 23.8 |
| *Secondary education* | 135 | 33.8 |
| *Tertiary education* | 146 | 36.5 |
| ***Marital Status*** |  |  |
| *Unmarried* | 180 | 45.0 |
| *Married* | 220 | 55.0 |
| ***Religion*** |  |  |
| *Christian* | 277 | 69.3 |
| *Islam* | 123 | 30.8 |
| ***Present Living Condition*** |  |  |
| *Living alone* | 74 | 18.5 |
| *Living with others* | 326 | 81.5 |
| ***Year of Diagnosis*** |  |  |
| *1997-2004* | 18 | 4.5 |
| *2005-2012* | 275 | 68.8 |
| *2013 and above* | 107 | 26.8 |
| ***Age at diagnosis*** |  |  |
| *27-37* | 57 | 14.2 |
| *38-48* | 233 | 58.3 |
| *49-59* | 90 | 22.5 |
| *60-70* | 20 | 5.0 |
| ***CD4 counts cells/mm<sup>3</sup>*** | 474.7±260.9 | 11-1675 |
| *Stage 1 (≥500 cells/µL)* | 169 | 42.3 |
| *Stage 2 (200-500 cells/µL)* | 179 | 44.8 |
| *Stage 3 (≤ 200 cells/µL)* | 52 | 13.0 |

### Clinical characteristics

According to the CDC’s staging system (Schneider et al., 2008), the largest percentage of participants were considered stage 2 (44.8%); followed by stage 1 (42.0%), and stage 3(12.8%). The mean (±S.D.) CD4 counts cells/mm^3^ was 434.9±260.2 with a range of 11-1675mm^3^. (Table 1)

### Psychiatric disorders and sleep quality

About 26% (95% CI: 21%–30%) of the participants had major depressive disorder, while 17.5% (95% CI: 14%– 21%) had anxiety disorder. Only 20.4% of participants with major depressive disorder and 24.3% of those with anxiety disorders had ever received biomedical specialist care. The prevalence of poor sleep quality was 29.3% (95% CI: 25%–34%), bipolar disorder was 3.3% (95% CI: 2.0%-5.0%), and mania was 1.0% (95% CI: 0%-2.0%). (Table 2).

**Table 2:** Prevalence of mental disorders among OPLHIV.

| *Variable* | *Mean (S.D)* | *Range* |
|---|---|---|
| ***Psychiatry morbidity*** | **12-month (n=400)** | **(%)** |
| ***Major depressive disorder*** |  |  |
| *No depressive disorder* | 297 | 74.2 |
| *Depressive disorder* | 103 | 25.8 |
| ***Ever received depression treatment (n=103)*** |  |  |
| *Yes* | 21 | 20.4 |
| *No* | 82 | 79.6 |
| ***Anxiety disorder*** |  |  |
| *No anxiety disorder* | 330 | 82.5 |
| *Anxiety disorder* | 70 | 17.5 |
| ***Ever received anxiety treatment (n= 70)*** |  |  |
| *Yes* | 17 | 24.3 |
| *No* | 53 | 75.7 |
| ***Sleep quality*** |  |  |
| *Poor sleep quality* | 117 | 29.3 |
| *Good sleep quality* | 283 | 70.7 |
| ***Mania disorder*** |  |  |
| *Yes* | 4 | 1.0 |
| *No* | 396 | 99.0 |
| ***Bipolar disorder*** |  |  |
| *Yes* | 13 | 3.3 |
| *No* | 387 | 96.7 |

### Virological suppression

Data were abstracted for viral load analyses, including 369 (92.3%) in 2019, 366 (91.5%) in 2020, 387 (96.8%) in 2021, 378 (94.5%) in 2022 and 368 (92%) in 2023.

Compared with the reference year (2019), the odds of viral suppression were significantly higher in 2020 (OR = 2.27, 95% CI: 1.26–4.12; p = 0.007), 2021 (OR = 2.17, 95% CI: 1.22–3.88; p = 0.009), and 2023 (OR = 3.43, 95% CI: 1.70–6.92; p < 0.001). Although the odds of viral suppression were higher in 2022 (OR = 1.66, 95% CI: 0.98– 2.81), the difference was not statistically significant (p = 0.061). Year was additionally modelled as a continuous variable, which shows a significant overall temporal trend, with the odds of viral suppression increasing by approximately 25% per year over the study period (OR = 1.25, 95% CI: 1.07–1.47, p = 0.006).

**Table 3:** Generalized Estimating Equation (GEE) Analysis of Viral Load Suppression by Year and trend analysis.

| *Viral Load* | *Viral suppression n (%)* | *Odds ratio* | *95% CI* | *P-value* | *Std.Error* | *Coefficient (B)* |
|---|---|---|---|---|---|---|
| *2019* | 330 (90.4) | Ref |  |  |  |  |
| *2020* | 345 (95.0) | 2.273 | 1.256-4.116 | 0.007 | 0.303 | 0.821 |
| *2021* | 360 (94.2) | 2.173 | 1.218-3.877 | 0.009 | 0.295 | 0.776 |
| *2022* | 343 (93.5) | 1.656 | 0.976-2.808 | 0.061 | 0.269 | 0.504 |
| *2023* | 355 (96.5) | 3.429 | 1.699-6.918 | <.001 | 0.358 | 1.232 |
| *Year trend* |  | 0.799 | 0.680-0.938 | 0.006 | 0.082 | -0.225 |

\*Variables significant at <.05

#### Clinical and demographic correlates of mental disorders

In the adjusted logistic regression models, sustained viral suppression was not significantly associated with current depressive symptoms (OR = 0.83, 95% CI: 0.51–1.34, p = 0.438) or anxiety symptoms (OR = 1.07, 95% CI: 0.61– 1.87, p = 0.811). However, sustained suppression was significantly associated with higher odds of good sleep quality (OR = 1.69, 95% CI: 1.07–2.66, p = 0.025).

Baseline CD4 count, age, marital status, and educational level were not significantly associated with any of the psychiatric outcomes. Female sex at birth was independently associated with higher odds of depressive symptoms (OR = 2.04, 95% CI: 1.16–3.60, p = 0.013) but showed no significant relationship with anxiety or poor sleep quality. (Table 4).

**Table 4:** Clinical and demographic correlates of mental disorders.

| **Correlates** | **Major depressive disorder**<br>**Odds ratio (95% CI), p-value** | **Anxiety disorder**<br>**Odds ratio (95% CI), p-value** | **Good Sleep quality**<br>**Odds ratio (95% CI), p-value** |
|---|---|---|---|
| Sustained viral suppression | 0.827 (0.512-1.337), 0.438 | 1.071 (0.612-1.873), 0.811 | 1.687 (1.067-2.661), 0.025* |
| CD4 count | 1.000 (0.999-1.001), 0.421 | 1.000 (0.999-1.001), 0.810 | 1.000 (0.999-1.001), 0.557 |
| Age | 1.027 (0.990-1.064), 0.155 | 1.000 (0.958-1.044), 1.000 | 0.981 (0.947-1.015), 0.268 |
| Sex at birth | 2.044 (1.160-3.602), 0.013* | 1.020 (0.541-1.923), 0.951 | 0.891 (0.525-1.512), 0.668 |
| Marital status | 1.302 (0.789-2.148), 0.302 | 1.147 (0.640-2.056), 0.644 | 0.928 (0.572-1.507), 0.764 |
| Educational status | 1.025 (0.972-1.080), 0.362 | 1.041 (0.979-1.107), 0.202 | 1.001 (0.952-1.053), 0.964 |

\*Variables significant at <.05, Major depressive and anxiety disorders were assessed using the WHO WMH-CIDI and coded as binary variables (1 = diagnosis present; 0 = diagnosis absent). Sleep quality was assessed using PSQI and coded as 1 = good sleep quality and 0 = poor sleep quality.

## Discussion

In this study among OPLHIV in Nigeria, which sought to examine the relationship of viral suppression, mental health, and sleep quality, sustained virologic suppression was not associated with major depressive disorder and anxiety disorder, suggesting they may be influenced by factors other than virologic status, but was associated with sleep quality.

Viral suppression remained consistently high across the study period, with evidence of a modest but statistically significant improvement over time. This pattern is consistent with reports from longitudinal HIV cohorts, where sustained engagement in HIV care, improved adherence, and advances in antiretroviral therapy have been associated with better virological outcomes over time[35, 36]. However, sustained viral suppression was not independently associated with depressive or anxiety symptoms. This aligns with other evidence from sub-Saharan Africa and other low- and middle-income settings showing that depression and anxiety are often driven more by psychosocial determinants including stigma, economic hardship, and reduced social support than by viral load measurement[37, 38]. A systematic review and meta-analysis across African populations found that approximately one-third of people with HIV experience depressive symptoms, predicted by female sex, stigma, and poor social support but not virological markers [37]. These findings, together with our observation of the relationship between depression and sustained viral suppression, support the view that psychiatric morbidity in older adults with HIV is multidimensional and not solely a result of the virological level.

In contrast, sustained viral suppression was significantly associated with better sleep, indicating a reduced likelihood of poor sleep. This finding is important, as sleep disturbance remains common but under-addressed among PLHIV. A meta-analysis conducted by Lee et.al (2024) on the prevalence and moderating factors of sleep disturbances in people with HIV estimated self-reported sleep problems in about 58% of people with HIV, compared with 13–30% in the general population[39, 40]. Viral suppression may improve sleep through reduced systemic and brain inflammation, lower symptom burden, and better overall functioning[41, 42]. Inflammatory markers such as IL-1β, IL-6, and TNF-α, have been linked to sleep dysregulation among people with HIV[42].

Female sex at birth was independently associated with higher odds of depressive symptoms, even after adjusting for sustained viral suppression, baseline CD4 count, and other sociodemographic co-variates. This is consistent with findings from previous studies showing a high burden of depression among women living with HIV in sub-Saharan Africa[43, 44]. A recent systematic review reported depression prevalence of 5.9% to 61% among women living with HIV in Africa, with intimate partner violence, stigma, poverty, unemployment, and social isolation as major predictors[43]. The persistence of this sex at birth association, after adjustment for other factors, suggests that depression risk is driven more by unmeasured factors, which could include structural and psychosocial variables, than by clinical HIV markers. Previous studies have linked low CD4 counts and high viral loads to neuropsychiatric symptoms through HIV-associated neuroinflammation[45]. However, in the current ART era, viral suppression may also reflect treatment adherence, sustained engagement in care, and overall disease stability, all of which are influenced by psychosocial factors. Lowther et al., in a systematic review of persistent psychological symptoms and perceived stigma among PLHIV on ART, reported a high burden of mental health disorders despite effective ART[46]. Other studies have also shown a bidirectional relationship where depression may reduce the likelihood of viral suppression[47]. These findings suggest a complex relationship between psychiatric morbidity and HIV disease markers in older adults.

In this cohort of older adults, psychiatric morbidity was not associated with sustained viral suppression. This finding suggests that, among OPLHIV, psychiatric morbidity may be driven more by psychosocial and aging-related factors than by markers of HIV disease control. Social isolation, chronic inflammation associated with aging and the cumulative psychosocial burden of long-term survival with HIV may contribute to psychosocial distress in this population[48, 49]. Similarly, the absence of an association between baseline CD4 count and current psychiatric morbidity may reflect the long term clinical stability achieved with ART[50, 51] alongside adaptive coping developed over years of living with HIV[17],52] and long-term engagement in care[51]. The low proportion of participants with depression (5.3%) and anxiety (4.3%) who had ever received biomedical specialist care may also help explain why psychiatric morbidity persisted despite successful HIV disease control in this cohort, highlighting an important gap between HIV treatment and mental healthcare.

Similarly, the high prevalence of poor sleep alongside depression points to the neuropsychiatric burden of aging with HIV. Although poor sleep can occur as a side effect of some ART regimens (e.g., integrase inhibitors), its lack of association with viral load suggests that sleep disturbance may reflect psychological distress, comorbidity, physiological ageing and health related concern rather than HIV disease activity[53–56].

## Conclusion

Prior viral load suppression did not correspond with current mental health status, suggesting that psychiatric morbidity in OPLHIV is influenced by factors beyond virological and immunological status. This supports the need for a broader, biopsychosocial approach to geriatric HIV care, where routine screening for depression, anxiety, and poor sleep is integrated into routine clinical care, regardless of viral suppression.

## Data Availability

The data used in this study is not publicly available but may be made available to qualified researchers on reasonable request from the corresponding author.

## Acknowledgments

We thank the study participants for their time and valuable contributions to this research. We also acknowledge the staff at the HIV clinic, Infectious Disease Institute, College of Medicine, University of Ibadan for their support with participant recruitment and data collection. We are grateful to the record staff of IDI for their assistance with clinic records and administrative approvals granted by the AIDS Prevention Initiative in Nigeria (APIN) in Abuja and Ibadan.

## Limitation

This study has several limitations. The retrospective design does not allow investigation of the temporal relationship between psychiatric symptoms and viral suppression. Since psychiatric symptoms were measured at only one point in time, it was not possible to determine whether they developed before or after changes in viral suppression. As a result, the associations observed should not be interpreted as causal relationship. Future longitudinal studies that assess both mental health and HIV treatment outcomes over time would provide a better understanding of how these factors influence one another.

## Declarations

### Ethical approval

This study was performed in line with the principles of the Declaration of Helsinki. Approval was granted by the joint University of Ibadan/University College Hospital Ethical Review Committee (UI/EC/22/0411). Administrative approvals were granted by the AIDS Prevention Initiative in Nigeria (APIN) in Abuja and Ibadan.

### Informed consent

We provided every participant with an informational leaflet detailing the study’s objectives. Additionally, written informed consent was obtained from all individual participants included in the study. Before proceeding with the assessment, the participants were made to understand their right to withdraw from the study at any point which would not at any time affect the treatment they are receiving at the HIV centre.

## Author Contribution

Conceptualization: Olufisayo Elugbadebo, Bibilola D. Oladeji

Methodology: Olufisayo Elugbadebo, Bibilola D. Oladeji

Formal analysis and investigation: Olufisayo Elugbadebo, Chiamaka Egbuna, Lisa Kuhns

Writing-original draft preparation: Olufisayo Elugbadebo, Akintade Faithful, Chiamaka Egbuna

Writing-review and editing: Olufisayo Elugbadebo, Akintade Faithful, Chiamaka Egbuna, Lisa Kuhns, Bibilola D. Oladeji, Lauren Beach, Olayinka Omigbodun, Philip B. Gorelick

Funding acquisition: Philip B. Gorelick, Olayinka Omigbodun

Resources: Olutosin Awolude

Supervision: Bibilola D. Oladeji, Lisa Kuhns, Lauren Beach

